# Discovering Monogenic Patients with a Confirmed Molecular Diagnosis in Millions of Clinical Notes with MonoMiner

**DOI:** 10.1101/2021.07.05.21259995

**Authors:** David Wei Wu, Jonathan A. Bernstein, Gill Bejerano

**Author notes:** **Corresponding Author** Gill Bejerano, Stanford University, Stanford, CA 94305. Broad Institute of MIT and Harvard, Cambridge, MA, USA.

## Abstract

**Purpose:** Cohort building is a powerful foundation for improving clinical care, performing research, clinical trial recruitment, and many other applications. We set out to build a cohort of all patients with monogenic conditions who have received a definitive causal gene diagnosis in a 3 million patient hospital system.

**Methods:** We define a subset of half (4,461) of OMIM curated diseases for which at least one monogenic causal gene is definitively known. We then introduce MonoMiner, a natural language processing framework to identify molecularly confirmed monogenic patients from free-text clinical notes.

**Results:** We show that ICD-10-CM codes cover only a fraction of known monogenic diseases, and even where available, code-based patient retrieval offers 0.12 precision. Searching by causal gene symbol offers great recall but an even worse 0.09 precision. MonoMiner achieves 7-9 times higher precision (0.82), with 0.88 precision on disease diagnosis alone, tagging 4,259 patients with 560 monogenic diseases and 534 causal genes, at 0.48 recall.

**Conclusion:** MonoMiner enables the discovery of a large, high-precision cohort of monogenic disease patients with an established molecular diagnosis, empowering numerous downstream uses. Because it relies only on clinical notes, MonoMiner is highly portable, and its approach is adaptable to other domains and languages.

## Introduction

Medical cohort building, the process of collecting one or more sets of patients with a set of common prescribed attributes, is a foundational activity in a healthcare system striving to improve current practices and meet new challenges^1–3^. Careful analysis of a set of related patients can inform and improve diagnosis and clinical care^4,5^, aid in clinical trial recruitment^6,7^, provide the basis for answering research questions^8,9^, enable the development of machine learning tools for automated patient care^10,11^, and in general facilitate the implementation of evidence based medicine.

The study and treatment of genetic diseases, and specifically monogenic disorders, is an especially attractive area to utilize patient cohorts. Each year, approximately 7 million infants (5.3%) worldwide are born with a serious rare genetic disorder^12^. Included in this group of diseases are thousands of monogenic diseases caused by variants in a single gene^13^. While individually rare, it is estimated that around 4% of the entire human population is affected by monogenic diseases in aggregate^13,14^. Since many monogenic diseases are rare, it both difficult to establish their genetic basis^15^ and difficult to recognize them in the clinic by physicians who may lack domain expertise^16^. Furthermore, monogenic diseases can have similar presentations to other diseases^17,18^ and patients of the same monogenic disease can have a wide range of different phenotypes^19,20^. Therefore, having access to cohorts of molecularly confirmed monogenic disease patients has the potential to greatly improve the diagnosis and treatment of monogenic disorders, and improve research on these diseases.

Here, we show that the current structure of our EMR systems makes identifying monogenic disease patients difficult, by assessing the performance of both International Classification of Diseases 10 Clinical Modification (ICD-10-CM) code^21^ lookup and HUGO (Human Genome Organization) Gene Nomenclature Committee (HGNC) gene symbol^22^ string search. To improve monogenic disease patient cohort identification, we (1) curate a database of monogenic diseases and their causative genes and (2) introduce MonoMiner, a natural language processing (NLP) tool to automatically identify molecularly confirmed monogenic patients from free-text clinical notes.

## Materials and Methods

### Stanford Medical Record Data

We used de-identified patient medical records from the Stanford Research Repository (STARR) database, provided by Stanford Medicine^23^. STARR is a clinical data warehouse following the Observational Medical Outcomes Partnership (OMOP) Common Data Model (CDM)^24^. It contains Epic^25^ data snapshots from Stanford Health Care, Stanford Children’s Health, the University Healthcare Alliance, and Packard Children’s Health Alliance. All patient data were obtained and processed under protocols approved by Stanford Medicine.

### Defining a Monogenic Disease Knowledgebase

To create a set of diseases with at least one established monogenic cause, we used the Dec 28, 2020 snapshot of the Online Mendelian Inheritance in Man (OMIM), a database containing relationships between genetic diseases and genes^26^. Since OMIM contains both monogenic and non-monogenic diseases, its entries were filtered to establish a monogenic disease knowledgebase. Briefly, OMIM diseases IDs were subset to those annotated to have at least one known molecular basis. To be classified as monogenic, an OMIM disease ID must have been annotated by OMIM to (1) have a mendelian mode of inheritance, (2) have at least one relationship with a causative gene that is not provisional, (3) not be a susceptibility to a disease, (4) not be a nondisease (defined by OMIM to be mostly conditions that lead to abnormal laboratory test values, such as OMIM:229800, representing “Fructosuria, Essential”), and (5) not be a chromosome deletion or duplication syndrome (**Fig 1, Supplementary Methods**). Each monogenic disease ID was then mapped to its causative OMIM gene ID(s) via OMIM-designated disease-gene relationships. Furthermore, each OMIM gene was tagged with its OMIM-designated HUGO (Human Genome Organization) Gene Nomenclature Committee (HGNC) gene symbol^22^.

**Figure 1.**
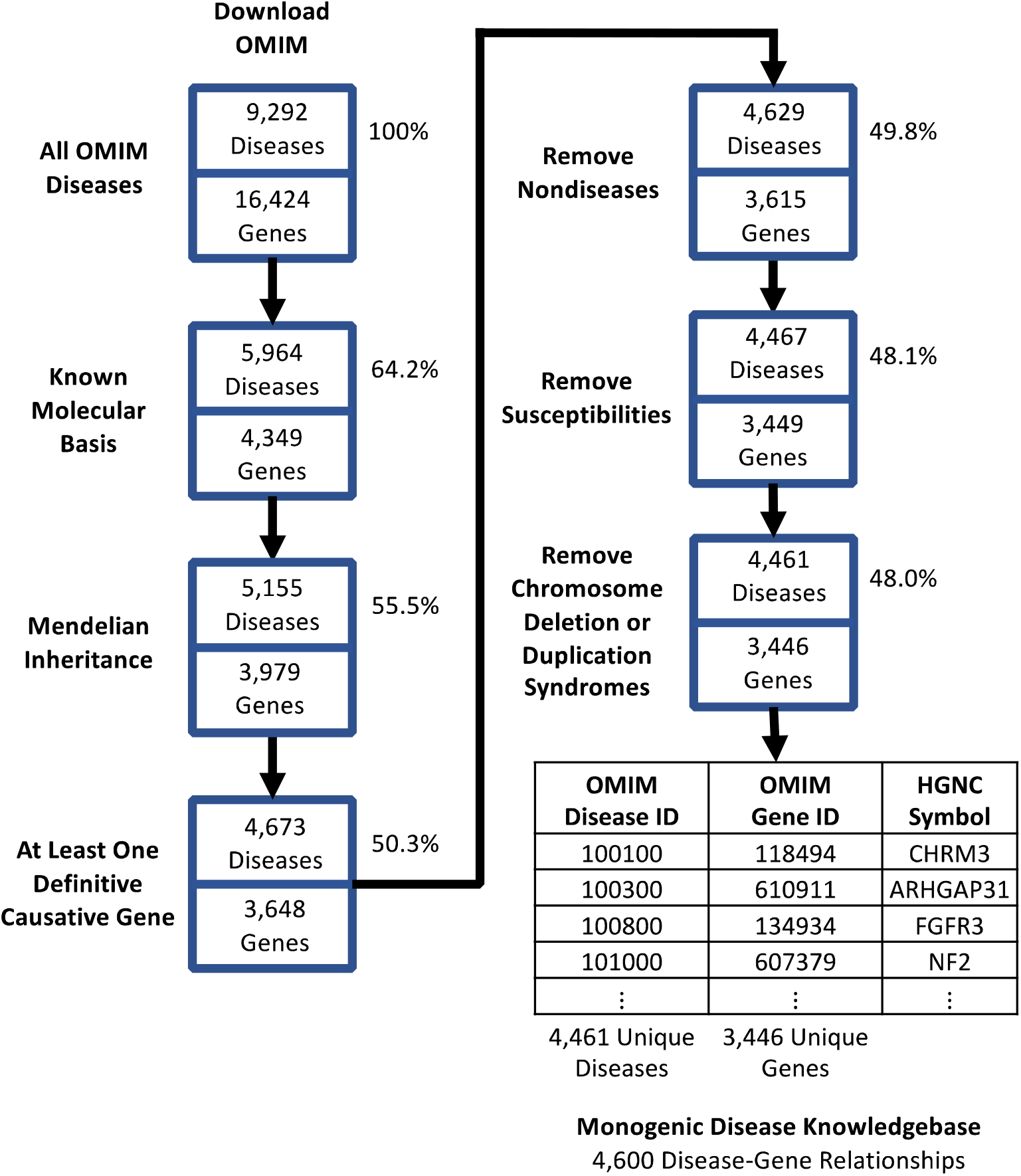
Creating a Monogenic Subset of OMIM. Set of filters to create our monogenic disease knowledgebase. All OMIM diseases and their associated genes are passed through 6 filters. The end result is a subset of OMIM diseases with at least one known monogenic causal gene.

### Mapping Monogenic Disease to ICD-9-CM and ICD-10-CM Codes

The International Classification of Diseases^27^ (ICD) is a comprehensive disease classification system used worldwide. OMIM provides mappings between OMIM diseases and ICD codes. To find monogenic patients using ICD codes, we obtained all OMIM-designated ICD-9 Clinical Modification^28^ (ICD-9-CM) and ICD-10 Clinical Modification^31^ (ICD-10-CM) codes relating to our monogenic subset of OMIM **(Fig 2)**.

**Figure 2.**
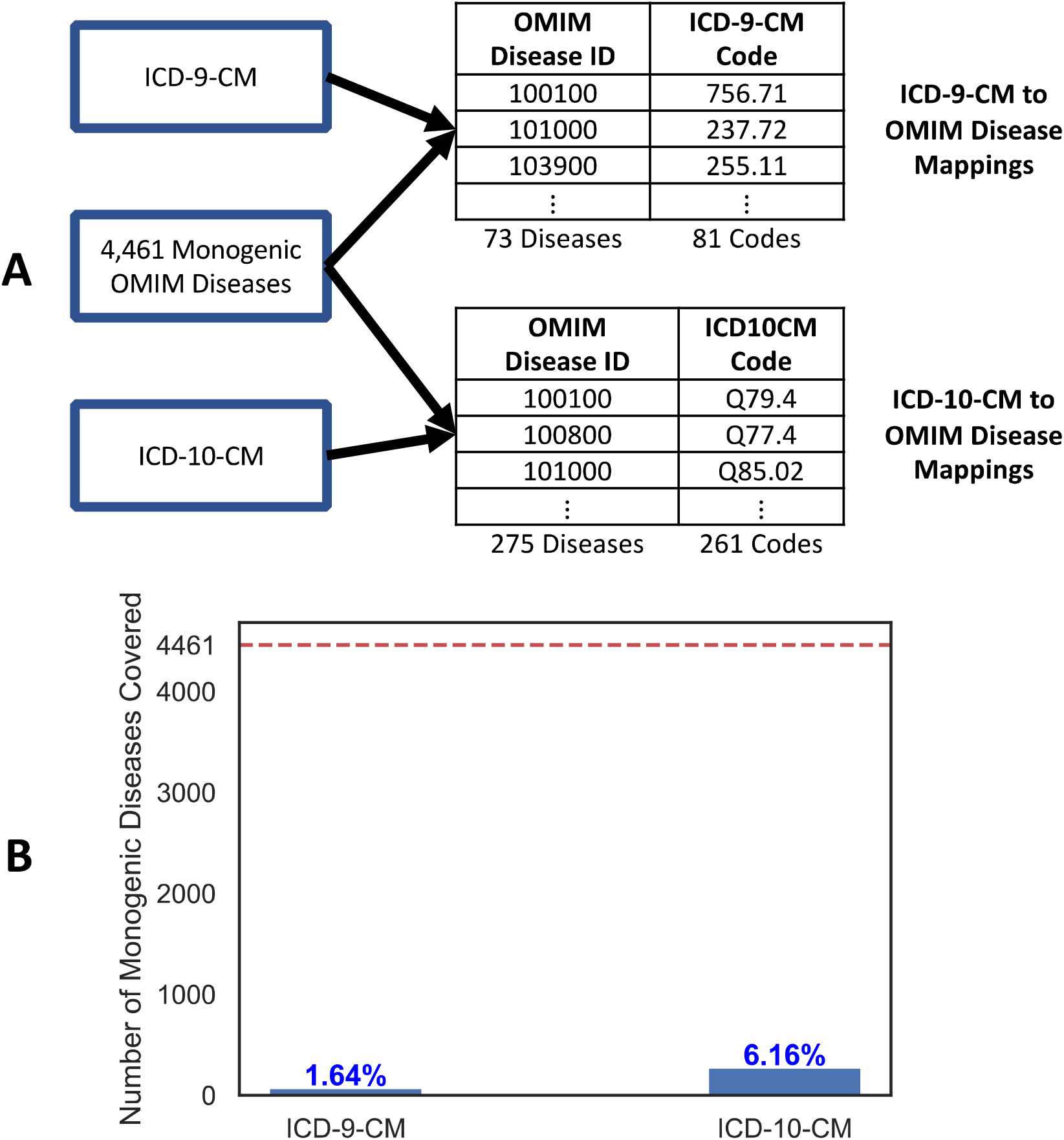
ICD Coverage of Monogenic Diseases. **(a)** The construction of both ICD-9-CM and ICD-10-CM mappings to monogenic OMIM diseases, provided by OMIM. **(b)** While ICD-10-CM improves on ICD-9-CM, both sets greatly lack in monogenic disease coverage.

### Querying Monogenic Disease Patients with ICD-10-CM Codes

To retrieve monogenic disease patients using ICD-10-CM, the STARR “Condition Occurrence” table, containing (patient, ICD-10-CM code) assignments, was queried for all patients who were annotated with one or more ICD-10-CM codes in our ICD-10-CM monogenic disease knowledgebase. The resulting list of (patient, ICD-10-CM code) pairs was converted to a list of (patient, possible OMIM disease ID) pairs using the ICD-10-CM monogenic disease knowledgebase. When an ICD-10-CM code mapped to more than one OMIM disease, we assigned the patient with all possible OMIM diseases.

### Querying Monogenic Disease Patients with Gene Symbol String Search

The HGNC^22^ is a worldwide authority that assigns unique and meaningful symbols to all genes in the human genome, allowing for unambiguous scientific communication. To mimic a physician manually searching for monogenic disease patients in their electronic health record (EHR) system, we searched patient notes for the HGNC gene symbol of each gene in the monogenic disease knowledgebase. Briefly, patient notes were tokenized into words by splitting on spaces, and a patient was annotated with possible OMIM disease ID *d* if their notes’ words contained the HGNC gene symbol for a causative gene of *d*. If a gene symbol mapped to more than one OMIM gene, we assigned the patient with all possible OMIM genes.

### Defining Disease and Gene Synonym Knowledgebases

Physicians use a variety of different terms to denote a particular monogenic disease in clinical notes. For example, “prune belly syndrome” and “eagle barrett syndrome” both refer to the OMIM disease OMIM:100100. Similarly, a variety of terms are used to refer to a single gene. For example, “CHRM3” and “EGBRS” both refer to the OMIM gene OMIM:118494. MonoMiner must have the capability to recognize the diverse set of synonyms for both diseases and genes. To this end, two knowledgebases were created. The disease synonym knowledgebase maps each monogenic OMIM disease ID to its synonyms. Similarly, the gene synonym knowledgebase maps each OMIM gene ID to its synonyms.

To create the disease synonym knowledgebase, each monogenic OMIM disease ID was mapped to its set of synonyms using both OMIM annotations and the Unified Medical Language System (UMLS) 2020AB metathesaurus^29^, a biomedical thesaurus linking synonyms of the same underlying concept from nearly 200 distinct vocabularies, including OMIM (**Fig 3a, Supplementary Methods**).

**Figure 3.**
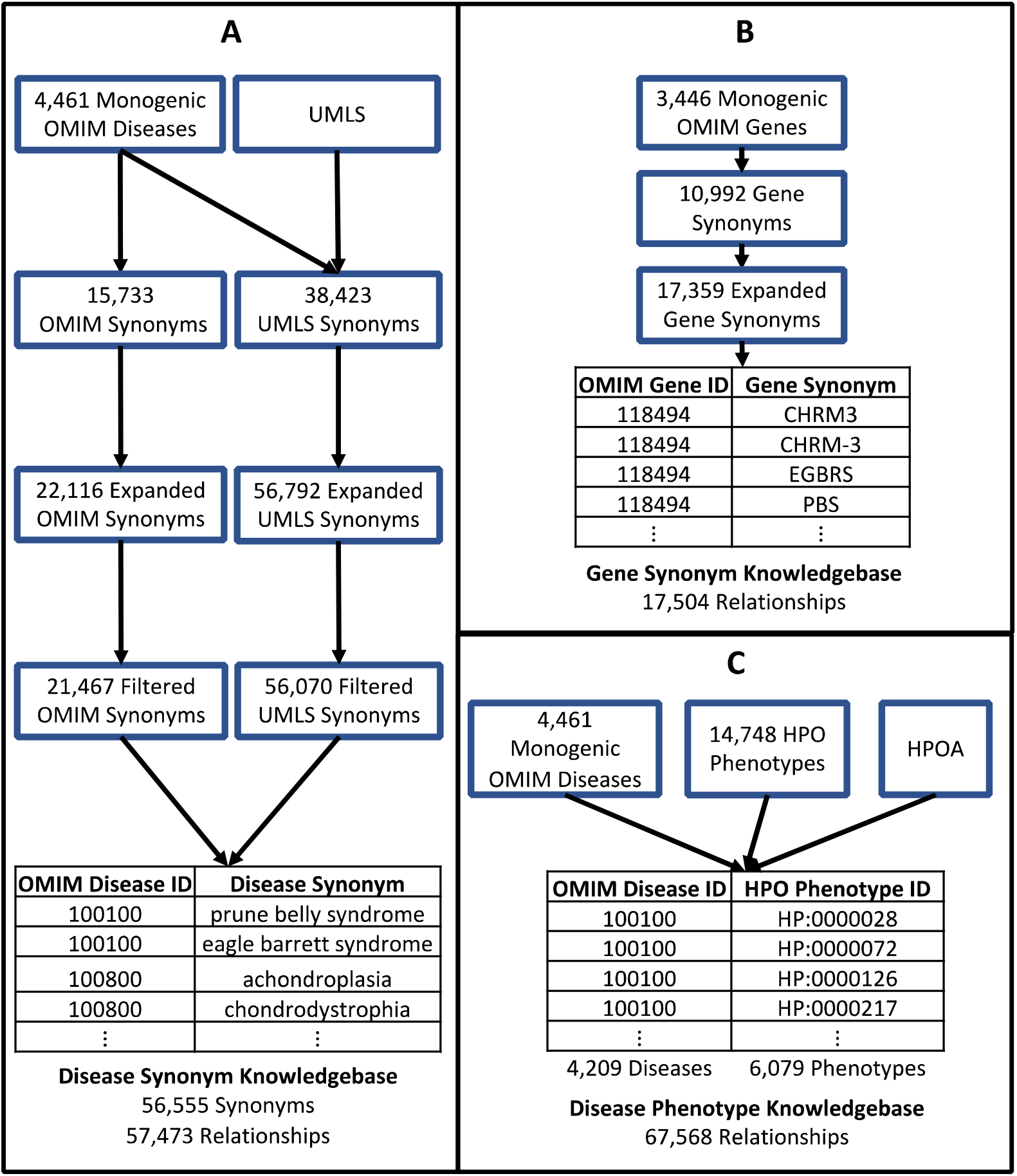
Creation of MonoMiner Knowledgebases. **(a)** Synonyms for diseases in the monogenic disease knowledgebase were obtained from both OMIM and UMLS. These sets were further expanded by removing punctuation from punctuation-containing synonyms. Next, synonyms likely to be nonspecific were excluded. The remaining synonyms were merged and mapped to their respective OMIM diseases. **(b)** The set of OMIM-designated gene synonyms for each monogenic disease gene were obtained and expanded by the addition of dashes (*e*.*g*. TBX-1), a practice we observed often in our doctor’s notes. **(c)** HPO-A provided us mappings between each OMIM monogenic disease and its associated HPO phenotypes.

To create the gene synonym knowledgebase, each OMIM gene ID in the monogenic disease knowledgebase was mapped to its OMIM-designated gene synonyms (**Fig 3b, Supplementary Methods**). We noticed that it was common practice for doctors to write a dash in gene symbols with an alphabetical prefix and numerical suffix. For example, “TBX1” is often written as “TBX-1”. Therefore, the gene synonym knowledgebase was expanded by the addition of mappings generated by inserting a dash between the alphabetical prefix and numerical suffix of such gene symbols.

### Defining a Disease Phenotype Knowledgebase

To refine its patient-disease identifications, MonoMiner compares patient phenotypes to known disease phenotypes (below). The Human Phenotype Ontology (HPO) (accessed March 27, 2020) was used to connect OMIM disease IDs to their known phenotypes. The HPO consists of two resources, an ontology hierarchically organizing human disease phenotypes and a set of phenotype to disease annotations (HPOA)^30^. All HPO phenotype IDs whose parent node is HP:0000118 (“phenotypic abnormality”) were mapped to our monogenic OMIM disease IDs using HPOA data **(Fig 3c)**.

### MonoMiner Overview

#### Preprocessing of Patient Free-Text Clinical Notes

To identify monogenic diagnoses, MonoMiner first splits patient notes into their component sentences and words **(Fig 4a)**. Briefly, a patient’s free-text clinical notes are processed twice, once with no modification to the notes and once with specific punctuation removed (**Supplementary Table S7**). In both passes, the delimiters “. “(period followed by space) and ““(four spaces/tab) are used to tokenize the notes into sentences. Periods that are part of honorifics (such as the period in “Dr.”) are ignored (**Supplementary Table S8**). Sentences are then split by space to obtain their component words. Finally, the resulting sentences and their component words from each pass are combined to be further analyzed.

**Figure 4.**
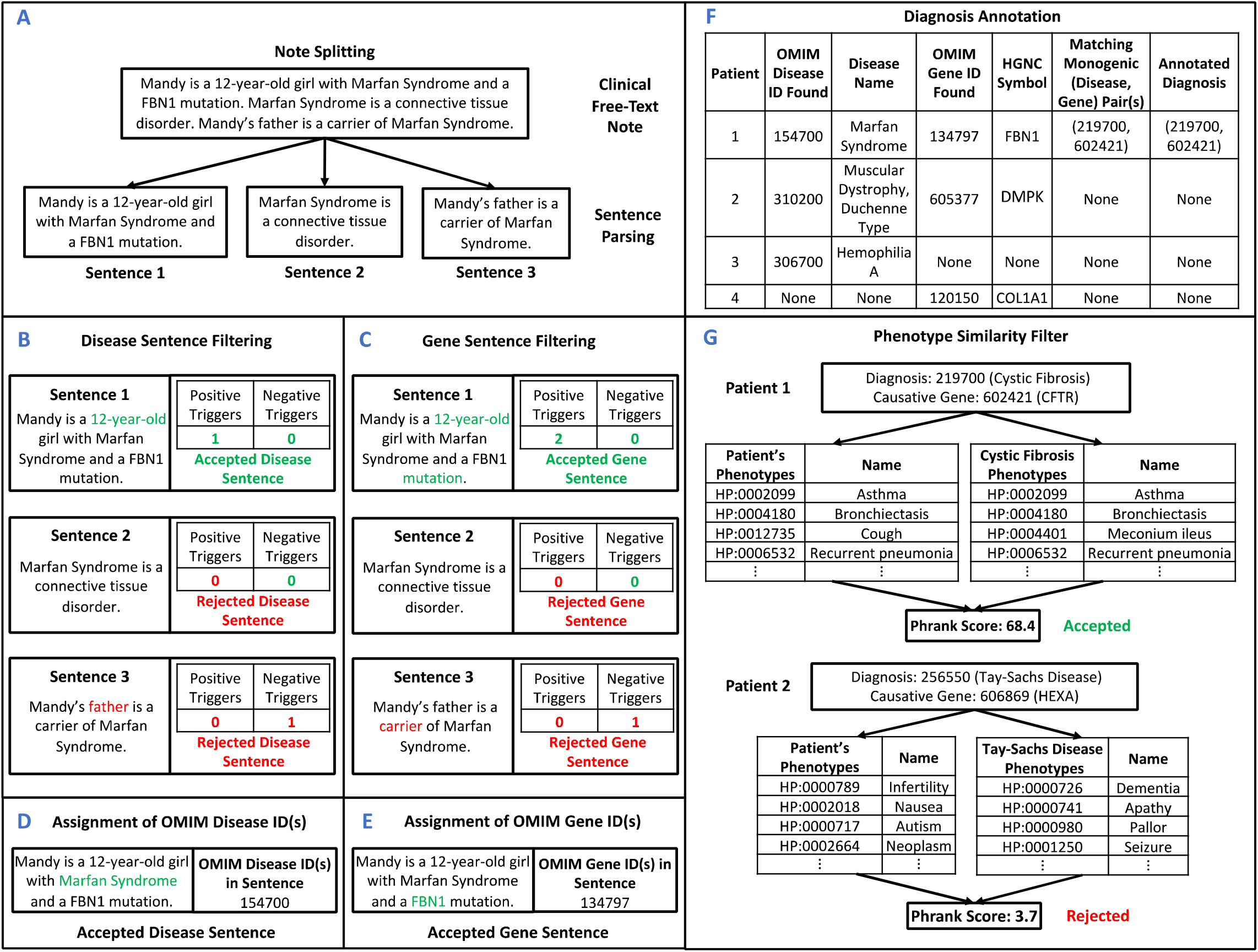
MonoMiner Pipeline. **(a)** We first split a patient’s notes into its component sentences **(b)** Sentence 1 is accepted as a candidate disease sentence because it (1) does not contain any negative disease trigger words and (2) contains a positive disease trigger word (“year-old”, green). Sentence 2 is not a candidate disease sentence because it does not contain any positive disease trigger words. Sentence 3 is not a candidate patient disease sentence because it contains a negative disease trigger word (“father”, red). **(c)** Sentence 1 is accepted as a candidate gene sentence because it (1) does not contain any negative gene trigger words, (2) it contains a positive patient trigger word (“year-old”, green), and (3) it contains a positive gene trigger word (“mutation”, green). Sentence 2 is not a candidate gene sentence because it does not contain any positive gene trigger words. Sentence 3 is not a candidate gene sentence because it contains a negative gene trigger word (“carrier”, red). **(d)** Candidate disease sentences are parsed for disease synonyms. **(e)** Candidate gene sentences are parsed for gene synonyms. **(f)** We then seek any matching (disease, gene) pair in our monogenic disease knowledgebase (Fig 1) and retain only patients with exactly one such pair. **(g)** Finally, patient phenotypes are automatically identified from the patient’s set of clinical notes using ClinPhen. Patient phenotypes are compared to possible disease phenotypes using Phrank. Only patients with some nontrivial amount of matching are retained.

#### Identification of Candidate Diagnostic Disease Sentences

Next, MonoMiner searches all sentences for candidate disease sentences **(Fig 4b)**. Candidate disease sentences are sentences potentially containing patient diagnosis information. Sentences are classified as candidate disease sentences based on the presence of positive trigger words (such as “diagnosis”), chosen to suggest the presence of relevant information, and the absence of negative trigger words (such as “normal”), chosen to indicate that the sentence is likely not relevant (**Supplementary Table S9, Supplementary Methods**). The sets of trigger words were determined during hyperparameter selection (see below).

#### Identification of Candidate Diagnostic Gene Sentences

MonoMiner also searches all sentences for candidate gene sentences **(Fig 4c)**. Similar to candidate disease sentences, candidate gene sentences are sentences potentially containing patient diagnostic genetic information. Identification of candidate gene sentences is analogous to identification of candidate disease sentences as described above. Sentences are classified as candidate gene sentences based on the presence of both positive patient trigger words (such as “confirm”) and positive gene trigger words (such as “variant”), and the absence of negative trigger words (such as “unknown”) (**Supplementary Table S10, Supplementary Methods**). Similar to candidate disease sentences, the sets of trigger words were determined during hyperparameter selection (see below). Of note, the same sentence can be both a candidate disease sentence and a candidate gene sentence (as in **Fig 4a**).

#### Assigning Monogenic OMIM Disease IDs to Patients

MonoMiner finds all synonyms in the disease synonym knowledgebase contained within a patient’s candidate disease sentences **(Fig 4d, Supplementary Methods)**. These disease synonyms are used to assign OMIM disease ID(s) to the patient via the disease synonym knowledgebase (**Fig 3a**). If a synonym mapped to more than one disease, the patient was tagged with all of them.

#### Assigning Monogenic OMIM Gene IDs to Patients

MonoMiner also finds all gene synonyms in the gene synonym knowledgebase contained within a patient’s candidate gene sentences **(Fig 4e, Supplementary Methods)**. These gene synonyms are used to assign the patient with OMIM gene ID(s) via the gene synonym knowledgebase (**Fig 3b**). If a synonym mapped to more than one gene, the patient was tagged with all of them.

#### Patient Diagnosis Assignment

At this stage, a patient has been assigned a set of zero or more candidate OMIM disease IDs from any of their candidate disease sentences across all their different notes and a similar set of zero or more candidate OMIM gene IDs. All (OMIM disease ID, OMIM gene ID) combinations from these two sets are generated, and the patient is annotated with any pair that is contained in our monogenic disease knowledgebase **(Fig 1)**. All other pairs are dropped. Only patients assigned exactly one (OMIM disease ID, OMIM gene ID) pair are kept **(Fig 4f)**.

#### Phenotype Similarity Filter

The previous step identifies patients with a possible monogenic disease diagnosis and associated causative gene as discussed in their notes **(Fig 4g)**. These patients are further filtered based on their clinical phenotypes. MonoMiner accomplishes this filtering by utilizing two existing algorithms, ClinPhen^31^ and Phrank^32^. ClinPhen is a natural language processing (NLP) method that identifies patient HPO phenotypes in clinical notes using a similar positive and negative trigger word approach to MonoMiner^31^. Phrank is an information theory-based framework to compute the similarity between any two sets of phenotypes^32^. Intuitively, patients whose phenotypes are not similar to the established phenotypes of their MonoMiner-assigned disease are discarded. Briefly, each patient has their top (see below) ClinPhen-ranked phenotypes identified. Then, the established phenotypes of their annotated OMIM disease are retrieved from the disease phenotype knowledgebase (**Fig 3c**). Finally, Phrank calculates the similarity between these two sets of phenotypes. Patient-disease assignments are discarded if their associated Phrank score is below a cutoff. The number of top ClinPhen-ranked phenotypes to identify per patient and the Phrank cutoff score were determined during hyperparameter selection (see below).

### MonoMiner Training Set

To select MonoMiner’s hyperparameters, we randomly selected a training set of 400,000 patients with clinical notes from the May 16th 2020 snapshot of STARR. All hyperparameters mentioned above were tuned to optimize performance on this training set.

Briefly, sets of candidate positive disease sentence trigger words, negative disease sentence trigger words, positive gene sentence trigger words, and negative gene sentence trigger words were manually curated by inspection of training set notes. Hyperparameter tuning began by initializing MonoMiner with no trigger words. Then, the single trigger word from any of the four classes of trigger words that most improved precision and recall was selected for inclusion in MonoMiner’s final trigger word sets. This process was repeated until the addition of trigger words did not result in an appreciable improvement in performance. At each stage, multiple values for the number of top ClinPhen phenotypes and Phrank cutoff score were considered, and the values that maximized performance on the last stage of tuning were used in the final version of MonoMiner (Supplementary Methods).

#### Assessing ICD-10-CM Code Search Per-Disease Precision

To determine the per-disease precision for ICD-10-CM code search, we created a test set of random patients from a hundred random monogenic diseases. Briefly, one hundred monogenic diseases with at least one ICD-10-CM annotated patient were randomly selected. For each sampled disease *d*, a patient was randomly selected for inclusion in the test set from the population of patients annotated by ICD-10-CM code with *d*.

Notes from each patient *p* in the test set with annotated disease *d* were manually reviewed, and *p* was classified for being “clinically diagnosed” and “molecularly confirmed.” To be “clinically diagnosed,” *p* must have been noted to have a diagnosis of *d*. In order to be classified as “molecularly confirmed,” *p* must have been “clinically diagnosed” and noted to have a variant in one of *d*’s causative genes not marked in the notes as “benign,” “likely benign,” or “of uncertain significance” according to American College of Medical Genetics and Genomics (ACMG) criteria^28^. Information regarding the exact nature of the variant (*e*.*g*. 76A>C or 83_84insTG) was not required to be noted. Precision was calculated as the fraction of test set that was “clinically diagnosed” and “molecularly confirmed”.

#### Assessing Gene Symbol String Search Per-Disease Precision

The process of determining the per-disease precision of gene symbol string search is analogous to that of ICD-10-CM code search. For a patient to be “clinically diagnosed”, the disease noted in their notes must have been one caused by the gene we searched for. For the patient to be “molecularly confirmed”, the molecular diagnosis noted in their notes must have been the same gene we searched for.

#### Assessing MonoMiner Per-Disease Precision

The process of determining the precision of MonoMiner is analogous to that of gene symbol string search. To ensure an unbiased estimate, any patient in MonoMiner’s training set was excluded from the test set.

An alternative per-patient precision estimate and a per-disease recall assessment are considered in the Discussion section.

#### Assessing Per-Patient Recall of All Methods

We curated one representative test set of random monogenic disease patients from the May 16, 2020 STARR snapshot to assess the recall of all three patient identification methods. First, we identified a broad set of potential monogenic patients as patients with the phrase “medical genetics” and not the term “perinatal” (an indicator that the note is about the patient’s unborn child) in at least one of their notes (Supplementary Methods). Individuals were randomly selected from this pool of 16,054 potential monogenic patients to have their notes manually reviewed for any disease-gene pair in the monogenic disease knowledgebase **(Fig 1)**. If such a disease-gene pair (*d, g*) was found and the patient (1) met the criteria of being “molecularly confirmed” for disease *d* and (2) had *g* as their underlying causative gene, they were annotated with (*d, g*) and added to the test set. This process was repeated until the test set reached the desired size. A total of 822 potential monogenic patients were manually inspected to create a recall test set of fifty. For each method, a patient *p* in the test set was classified as successfully “retrieved” if the respective method tagged them with *d*. For MonoMiner and gene symbol string search, *p*’s causative gene annotation must have also been *g*. Recall was calculated as the fraction of successfully retrieved patients.

## Results

The main goal of this work is to develop an efficient method to find monogenic disease patients with a confirmed molecular diagnosis. Our top priority is to minimize false positive identifications, or maximize precision, with a secondary consideration given to minimize false negatives, or maximize recall.

### Monogenic Disease Knowledgebase

Our copy of OMIM contains 9,292 unique diseases. Selecting for all diseases with at least one established monogenic causal gene results in 4,461 (48%) unique OMIM disease IDs caused, at least in part, by 3,446 unique human genes (see **Methods, Fig 1, Supplementary Table S1**)

### ICD Codes Have Poor Coverage of Monogenic Diseases

Using OMIM mappings, the final ICD-9-CM knowledgebase consisted of only 81 OMIM disease ID to ICD-9-CM code mappings covering 73 OMIM disease IDs and 81 ICD-9-CM codes **(Fig 2a, Supplementary Table S5)**. The final ICD-10-CM knowledgebase consisted of only 307 OMIM disease ID to ICD-10-CM code mappings covering 275 OMIM disease IDs and 261 ICD-9-CM codes **(Fig 2a, Supplementary Table S6)**.

Importantly, we observed just 1.64% (73 out of 4,461) monogenic disease coverage by ICD-9-CM codes. While ICD-10-CM codes increased monogenic disease coverage almost four-fold to 6.16% (275 out of 4,461), a large majority of monogenic diseases are not represented **(Fig 2b)**. Because the ICD code search space of diseases makes up a small fraction of the entire monogenic disease space, many monogenic disease patients cannot be retrieved with these codes (affecting recall, but not precision, below).

### ICD-10-CM Code Search Has Low Precision and Recall

We assessed both the precision and recall of using ICD-10-CM codes to retrieve patients with the 275 (6.16% of) monogenic diseases covered by ICD-10-CM. In total, this search returned 133,171 (4.3%) of 3,074,787 patients, spanning 261 unique diseases of 275 covered. Out of all patients in the precision test set, 35% had a clinical diagnosis that matched with their annotated disease (“Clinically Diagnosed”), but only 12% were documented with a genetic variant consistent with their disease (“Molecularly Confirmed”) **(Table 1, Supplementary Table S11)**. In the group of false positives, two common sources of error included annotating patients with an ICD-10-CM code of a disease being considered during a differential diagnosis and annotating patients with an ICD-10-CM code of a disease that a patient was undergoing a laboratory test for. In addition, two of every three patients (68%) in our recall set were not retrievable using ICD code search, as their OMIM disease was not mapped to any code, while recall itself was the complementary 0.32 **(Table 1, Supplementary Tables S6 and S14)**.

**Table 1.** Performance Comparison of Three Methods to Find Monogenic Patients with a Confirmed Casual Gene from Clinical Notes. ICD-10-CM covers a small fraction of monogenic diseases (**Fig 2**). It overpredicts these diseases, mostly covering more common diseases with only a small fraction of monogenic patients (**Table 3**). String search provides great recall at the cost of greatly overpredicting coverage over both patients and diseases (**Table 4**). MonoMiner greatly improves precision, which is critical for physician consultation and machine learning training alike (**Table 2**). Per-disease Monominer performance is 2.5-9 times better than ICD-10-CM and gene symbol string search. **Tables 2-4** strongly suggest that the gap would be even bigger for per-patient precision.

|  | ICD-10-CM<br>Code Search | Gene Symbol String<br>Search | MonoMiner |
| --- | --- | --- | --- |
| **Number of Diseases in Search Space**<br>(Percentage of All 4,461 monogenic diseases) | 275<br>(6.2%) | 4,461<br>(100%) | 4,209<br>(94.4%)* |
| **Number of Patients Retrieved**<br>(Percentage of Stanford 3,074,787 adult and pediatric patients) | 133,171<br>(4.3%) | 1,108,377<br>(36.0%) | 4,259<br>(0.1%) |
| **Number of Monogenic Diseases Hypothesized**<br>(Percentage of all 4,461 monogenic diseases) | 261<br>(5.9%) | 4,097<br>(91.8%) | 560<br>(12.6%) |
| **Number of Causative Genes Hypothesized**<br>(Percentage of all 3,446 monogenic genes) | N/A | 3,102<br>(90%) | 534<br>(15.5%) |
| **Clinically Diagnosed Per-Disease Precision** | 0.35 | 0.13 | 0.88 |
| **Molecularly Confirmed Per-Disease Precision** | 0.12 | 0.09 | 0.82 |
| **Per-Patient Recall** | 0.32 | 0.96 | 0.48 |

\*MonoMiner is currently unable to retrieve patients with 252 monogenic OMIM diseases because they have yet to be tagged with HPO phenotypes in HPOA. In practice, on the Nov 28 2020 STARR database, only 3 patients of 3 million searched were candidates for one of these diseases.

Analysis of the top 10 most frequently annotated ICD-10-CM terms (**Table 2**) further reveals ICD-10-CM’s weaknesses in retrieving monogenic disease patients. For disorders in which only a small subpopulation of affected patients are monogenic cases, such as hypercholesterolemia, ICD-10-CM codes do not have the resolution to distinguish between truly monogenic cases and cases caused by environmental or other factors. Additionally, ICD-10-CM codes can be nonspecific, representing a broad class of diseases. For example, four monogenic OMIM diseases, Lecithin:cholesterol acyltransferase deficiency (OMIM:245900), Familial Hypobetalipoproteinemia 1 (OMIM: 615558), Abetalipoproteinemia (OMIM: 200100), and Tangier Disease (OMIM:205400), all share the same OMIM-designated ICD-10-CM code, E78.6, which codes for the general term “Lipoprotein deficiency”.

**Table 2.** Top Ten Most Frequently Retrieved ICD-10-CM Codes.

| ICD-10-CM Code | ICD-10-CM Code Name | OMIM disease ID | OMIM Disease Name | Patients Retrieved | Percentage of Total Retrieved Patients | MedLine Plus<sup>38</sup> Frequency of OMIM Disease |
| --- | --- | --- | --- | --- | --- | --- |
| E78.2 | Mixed hyperlipidemia | 144010 | Hypercholesterolemia, Familial, 2 | 41,140 | 23.77% | 1 in 200 to 250 |
| E78.00 | Pure hypercholesterolemia, unspecified | 143890 | Hypercholesterolemia, Familial, 1 | 39,981 | 23.1% | 1 in 200 to 250 |
| E78.49 | Other hyperlipidemia | 144250 | Hyperlipidemia, Familial Combined, 3 | 5,687 | 3.29% | 1 per million |
| E80.6 | Other disorders of bilirubin metabolism | 237500 | Dubin-Johnson Syndrome | 5,448 | 3.15% | 1 in 1,300 in Iranian and Moroccan Jews, lower elsewhere |
| I49.8 | Other specified cardiac arrhythmias | 601144 | Brugada Syndrome 1 | 3,768 | 2.18% | 1 in 2,000 |
| Q23.1 | Congenital insufficiency of aortic valve | 109730 | Aortic Valve Disease 1 | 3,466 | 2% | N/A |
| E78.6 | Lipoprotein deficiency | 615558 | Hypobetalipoproteinemia, Familial, 1 | 3,079 | 1.78% | 1 in 1,000 to 3,000 |
|  |  | 200100 | Abetalipoproteinemia |  |  | More than 100 cases. |
|  |  | 205400 | Tangier Disease |  |  | 100 cases |
|  |  | 245900 | Lecithin:Cholesterol Acyltransferase Deficiency |  |  | 70 known cases |
| I27.0 | Primary pulmonary hypertension | 178600 | Pulmonary Hypertension, Primary, 1 | 2,726 | 1.58% | 1,000 new US cases a year |
| G56.00;<br>G56.0 | Carpal tunnel syndrome, unspecified upper limb; Carpal tunnel syndrome | 115430 | Carpal Tunnel Syndrome | 2,586 | 1.49% | 1% to 5% of the adult population |
| K74.69 | Other cirrhosis of liver | 215600 | Cirrhosis, Familial | 2,513 | 1.45% | N/A |

In conclusion, ICD-10-CM code search has both low precision, returning many patients who in fact do not have the disease they are annotated with, and low recall, missing the majority of real monogenic patients due to poor coverage of monogenic diseases.

### Gene Symbol String Search Achieves High Recall but Very Low Precision

Gene symbol string search resulted in the largest patient cohort, retrieving an unreasonable 36% of all Stanford patients (1,108,377 patients). These patients unrealistically spanned 92% (4,097) of all monogenic diseases and 90% (3,102) of all known monogenic disease genes. While nearly every patient (96%) in the recall test set was successfully retrieved (**Table 1, Supplementary Table S14**), precision was the lowest of all methods tested, with only 13% correctly “clinically diagnosed,” and only 9% correctly “molecularly confirmed” (**Table 1, Supplementary Table S12**).

The top ten most frequently annotated HGNC gene symbols all had alternative meanings leading to many false positives **(Table 3)**. For example “HR”, in addition to being the HGNC symbol of HR lysine demethylase and nuclear receptor corepressor (OMIM: 602302), the causative gene of Alopecia universalis (OMIM: 203655), Atrichia with papular lesions (OMIM: 209500), and Hypotrichosis 4 (OMIM:146550), is commonly used as an abbreviation of “heart rate”. Similar patterns were observed for “OTC” (over the counter), “CAD” (coronary artery disease), and others (**Table 3**).

**Table 3.** Top Ten Most Frequently Retrieved Genes with Gene Symbol String Search.

| HGNC Gene Symbol | OMIM Gene ID | OMIM Disease ID(s) | Disease Name | Patients Retrieved | Percentage of Total Retrieved Patients* | Disease Frequency (Medline Plus<sup>38</sup> and OrphaNet<sup>39</sup>) |
| --- | --- | --- | --- | --- | --- | --- |
| HR | 602302 | 146550 | Hypotrichosis 4 | 319,923 | 28.86% | N/A |
|  |  | 203655 | Alopecia Universalis Congenita |  |  | 1 to 5 in 10,000 |
|  |  | 209500 | Atrichia with Papular Lesions |  |  | N/A |
| OTC | 300461 | 311250 | Ornithine Transcarbamylase Deficiency | 282,388 | 25.48% | 1 in 14,000 to 77,000 |
| CAD | 114010 | 616457 | Developmental and Epileptic Encephalopathy 50 | 171,252 | 15.45% | Less than 1 in 1,000,000 |
| CP | 117700 | 604290 | Aceruloplasminemia | 166,888 | 15.06% | 1 in 2 million |
| KY | 605739 | 617114 | Myopathy, Myofibrillar, 7 | 135,618 | 12.24% | Less than 1 in 1,000,000 |
| WAS | 300392 | 300299 | Neutropenia, Severe Congenital, X-linked | 129,495 | 11.68% | 1 in 200,000 |
|  |  | 301000 | Wiskott-Aldrich Syndrome |  |  | 1 to 10 cases per million males |
|  |  | 313900 | Thrombocytopenia 1 |  |  | 1 to 10 cases per million males |
| FH | 136850 | 150800 | Hereditary Leiomyomatosis and Renal Cell Cancer | 111,355 | 10.05% | 300 families |
|  |  | 606812 | Fumarase Deficiency |  |  | 100 individuals |
| SI | 609845 | 222900 | Sucrase-Isomaltase Deficiency, Congenital | 108,088 | 9.75% | 1 in 5,000 Europeans |
| ACE | 106180 | 267430 | Renal Tubular Dysgenesis | 71,658 | 6.47% | N/A |
| C3 | 120700 | 613779 | Complement Component 3 Deficiency, Autosomal Recessive | 71,498 | 6.45% | Less than 1 in 1,000,000 |

\* The sum of this column exceeds 100% because patients frequently have multiple HGNC gene symbols in their clinical notes

Note also that because the recall of gene symbol string search is so high, looking back at the number of patients retrieved using ICD-10-CM codes (**Table 3**), it is clear the vast majority do not have a matching molecular diagnosis (*e*.*g*. 41,140 patients with the ICD-10-CM code for Familial Hypercholesterolemia 2, but only 176, or 233 times fewer, patients are found by gene symbol string search for LDLR, the causative gene of Familial Hypercholesterolemia 2).

### MonoMiner’s Disease Synonym Knowledgebase

As both ICD and gene symbol string search leave much to be desired in terms of precision-recall balance, we turn to set up the knowledgebases underlying our alternative method, named MonoMiner.

To establish MonoMiner’s disease synonym knowledgebase, we queried both OMIM and UMLS for disease names of the 4,461 monogenic OMIM disease IDs in the monogenic disease knowledgebase. A total of 15,733 and 38,423 disease synonyms were obtained from OMIM and UMLS, respectively. Expanding these sets of synonyms by the addition of synonyms stripped of punctuation (Supplementary Methods) resulted in 22,116 and 56,792 synonyms for OMIM and UMLS, respectively. Synonym filtering for nonspecific synonyms (Supplementary Methods) reduced OMIM synonyms to 21,467 and UMLS synonyms to 56,070. Finally, combining the synonyms from the two databases together yielded the final disease synonym knowledgebase, containing 57,473 OMIM disease ID to disease synonym mappings covering all 4,461 monogenic diseases in our monogenic disease knowledgebase **(Fig 3a, Supplementary Table S2)**.

### MonoMiner’s Gene Synonym Knowledgebase

To create MonoMiner’s gene synonym knowledgebase, all OMIM-designated gene symbols for the 3,446 genes in our monogenic disease knowledgebase were obtained, resulting in a set of 10,992 gene synonyms. Expanding these gene synonyms using dashes (Supplementary Methods) increased their number to 17,359. Mapping of these gene synonyms back to their respective OMIM gene IDs resulted in the final gene synonym knowledgebase, consisting of 17,504 unique (OMIM gene ID, gene synonym) mappings covering all 3,446 OMIM gene IDs in our monogenic disease knowledgebase **(Fig 3b, Supplementary Table S3)**.

### MonoMiner’s Disease Phenotype Knowledgebase

To associate monogenic diseases with their phenotypes, the entire HPO database, consisting of 14,748 phenotypes, was obtained. Next, all phenotypes belonging to the “phenotypic abnormality” class were mapped to their respective monogenic diseases in the monogenic disease knowledgebase using HPO-A annotations. This resulted in a disease phenotype knowledgebase with 67,568 OMIM disease ID to HPO phenotype ID mappings, covering 4,209 (94%) monogenic OMIM disease IDs and 6,079 HPO phenotypes **(Fig 3c, Supplementary Table S4)**.

### MonoMiner Identifies Accurate Patient Annotations with Moderate Recall

When run on the November 28, 2020 release of STARR, MonoMiner tags 4,259 patients with specific disease and causal gene diagnoses, spanning 560 unique diseases and 534 unique causative genes **(Supplementary Table S15)**. MonoMiner significantly outperforms both ICD-10-CM code search and gene symbol string search in per-disease precision, achieving 88% “clinically diagnosed” and 82% “molecularly confirmed” precision **(Table 1, Supplementary Table S13)**, with recall of 48% **(Table 1, Supplementary Table S14)**.

Unlike ICD-10-CM code search and gene symbol string search, MonoMiner’s 10 most frequently annotated diseases represent common monogenic disorders **(Table 4)**. Additionally, many top hits, like Cystic Fibrosis (OMIM:219700), have high penetrance and a dedicated treatment center at Stanford, potentially further explaining their enrichment in MonoMiner annotations.

**Table 4.** Top Ten Most Frequently Retrieved Diseases with MonoMiner.

| **OMIM Disease ID** | **Disease Name** | **HGNC Gene Symbol** | **Patients Retrieved** | **Proportion of Total Retrieved Patients** | **MedLine Plus<sup>38</sup> Frequency** |
| --- | --- | --- | --- | --- | --- |
| 219700 | Cystic Fibrosis | CFTR | 669 | 15.71% | 1 in 2,500 to 3,500 whites |
| 162200 | Neurofibromatosis, Type I | NF1 | 211 | 4.95% | 1 in 3,000 to 4,000 |
| 154700 | Marfan Syndrome | FBN1 | 178 | 4.18% | 1 in 5,000 |
| 154800 | Mastocytosis, Cutaneous | KIT | 137 | 3.22% | 1 per 10,000 to 20,000 |
| 175100 | Familial Adenomatous Polyposis 1 | APC | 98 | 2.30% | 1 in 7,000 to 1 in 22,000 |
| 235200 | Hemochromatosis, Type 1 | HFE | 91 | 2.14% | 1 in 350 |
| 151623 | Li-Fraumeni Syndrome | TP53 | 71 | 1.67% | 1 in 5,000 to 1 in 20,000 |
| 249100 | Familial Mediterranean Fever | MEFV | 70 | 1.64% | 1 in 200 to 1,000 in the Mediterranean population, lower elsewhere |
| 613254 | Tuberous Sclerosis 2 | TSC2 | 69 | 1.62% | 1 in 6,000 |
| 310200 | Muscular Dystrophy, Duchenne Type | DMD | 68 | 1.60% | 1 in 3,500 to 5,000 newborn males |

Error analysis was performed on the patients in the recall test set MonoMiner was unable to retrieve. While a variety of reasons were identified, the vast majority of patients were not successfully annotated by MonoMiner due to two main failure modes. The first occurred when all sentences that contained the patient’s molecular diagnosis information did not contain positive patient trigger words. An example of this type of sentence is “A deleterious variant in the FBN1 gene was found.” The second occurred when the synonyms that physicians used to refer to a disease were not found in the disease synonym knowledgebase. An example of this is physicians using the term “epilepsy” to refer to OMIM:617166, which represents the disease Developmental and Epileptic Encephalopathy 47.

## Discussion

Here we present MonoMiner, a Natural Language Processing (NLP) based approach to correctly identify monogenic patients with a gene-based diagnosis from free text clinical notes. We start by trying alternative methods, available in some Electronic Medical Record (EMR) systems today. We show that ICD-10-CM currently offers limited coverage of monogenic diseases, and that even when restricting to these diseases, finding molecular diagnosis via these codes has a precision worse than 1 in 8. String-based search, which is not even available in some EMR systems today, has very high recall, but at the price of many false positives. More than 1/3 of all 3 million pediatric or adult Stanford patients are unrealistically tagged as having a monogenic molecular diagnosis, and precision even at the level of diseases is lower than 1 in 11.

To fill this gap, MonoMiner develops an NLP approach. Positive and negative trigger words are optimized to search for indicative sentences, pairs of extracted monogenic diseases and genes are compared to an OMIM-derived knowledgebase of thousands of such known pairs, and two existing tools from the lab, ClinPhen^31^ and Phrank^32^, are used to automatically extract and rank patient phenotypes and ensure these patient phenotypes resemble the disease phenotypes as documented in OMIM. Using this approach, we obtain 0.88 precision for retrieving patients with a confirmed disease diagnosis and 0.82 precision for retrieving patients with a confirmed causal monogenic gene, at a recall of nearly ½.

The poor coverage of individual monogenic diseases in ICD-9-CM and ICD-10-CM is well known^34^. And while ICD-11 is set to improve this situation^27^, it is not here yet, and it will not cover all monogenic diseases even when deployed. As we show with ICD-10-CM use (**Table 2**), it will not be easy to accurately back-annotate pre-existing medical records with ICD-11 codes without resorting to NLP approaches.

We chose to evaluate the precision of all three methods on a per-disease basis, which means we randomly picked one patient per disease per method. MonoMiner is 7-9 times more precise than the other two methods according to this measure. Had we evaluated precision per-patient and chosen a random subset of patients from each approach, **Tables 2-4** make it clear that the precision gap will likely be even bigger in favor of MonoMiner. We cannot assess recall per disease because short of semi-manually annotating all 3 million patient records, we do not know how many diagnosed monogenic diseases are in our system.

Our per patient recall of 0.48 suggests MonoMiner can be improved on. Because correctly labeling every medical record is laborious, and takes in our experience on the order of minutes per case, it is hard to train rich models like deep learning approaches^35^ with little tagged data. Weak supervision^36^ offers an appealing approach, and our optimized rules (**Fig 4**) can serve as a set of labeling functions to introduce into such a model.

The 4,259 patients MonoMiner flags, covering 560 different monogenic diseases and 534 different causal genes, are a treasure trove. For example, a recent paper^37^ used external labeled data to build a specialized classifier to find molecularly diagnosed Familial Hypercholesterolemia (FH) cases in the Stanford system. Using their approach, the authors validated 66 such cases. MonoMiner impressively finds 64 molecularly diagnosed FH patients (while finding patients for many other diseases as well). Gene symbol search unrealistically flags a thousand FH cases and ICD-10-CM code search retrieves tens of thousands.

The approach we devise to glean information from the one place clinicians mostly use to converse with each other, the clinical notes, is very general. The strategy we use is also not exclusive to monogenic diseases. For example, removing the gene requirement and expanding the disease knowledgebase will allow one to find diagnosed patients for a variety of different diseases. As medical record systems continue to evolve, and until the standardization of structured data input and update is integrated into the clinicians daily work flow, Natural Language approaches such as MonoMiner remain the best strategies for organizing and mining patient records.

## Data Availability

This research used data or services provided by STARR, "STAnford medicine Research data Repository," a clinical data warehouse containing live Epic data from Stanford Health Care, the Stanford Children's Hospital, the University Healthcare Alliance and Packard Children's Health Alliance clinics and other auxiliary data from Hospital applications such as radiology PACS. STARR platform is developed and operated by Stanford Medicine Research IT team and is made possible by Stanford School of Medicine Research Office.

## Data Availability

MonoMiner will be available for download at the following website: https://bitbucket.org/bejerano/monominer.

## Acknowledgments

This research also used the Nero Platform to access STARR. Nero is developed and operated by Stanford Research Computing Center. STARR-OMOP on Nero is supported by Stanford Medicine Research IT team. The services provided by Nero Research Computing platform is made possible by Stanford School of Medicine Research Office and Stanford Research Computing Center.

We thank Nigam Shah, Somalee Datta, Priya Desai, Lingyao Yang, and all members of the Stanford Research Computing Center for data and resources availability, Kailas Vodrahalli, Cole Deisseroth, and Nik Caryotakis for technical advice, and all members of the Bejerano Lab for valuable discussions and project feedback.

## Author Information

Conceptualization: D.W.W., G.B.; Data curation: D.W.W.; Formal Analysis: D.W.W., J.A.B, G.B.; Funding acquisition: D.W.W., G.B.; Investigation: D.W.W.; Methodology: D.W.W., G.B.; Software: D.W.W.; Supervision: G.B.; Visualization: D.W.W.; Writing–original draft: D.W.W., G.B.; Writing–review and editing: D.W.W., J.A.B, G.B.

## Ethics Declaration

Access of deidentified patient electronic medical record data was approved under Stanford eProtocol 13655. This study was approved by Stanford IRB protocol #44566. All of our work was done on deidentified records, requiring no further patient consent.

